# Sam68 Exacerbates Pathological Cardiac Hypertrophy by Suppressing Cardiomyocyte Glucose Oxidation

**DOI:** 10.1101/2025.01.31.25321508

**Authors:** Junqing An, Chaoshan Han, Ying Jiang, Jiawei Shi, Huadong Li, Chenqi Wang, Jianrong Huang, Shiyue Xu, Nianguo Dong, Gangjian Qin

## Abstract

**Background:** Impaired energy metabolism in the heart is critical in the development of cardiac hypertrophy and failure. Src-associated in mitosis 68 kDa (Sam68) is a member of the signal transducer and activator of RNA (STAR) protein family, and its role in cardiac energy metabolism is undefined.

**Methods:** We assessed Sam68 expression in human myopathic and failing hearts. We also generated mice with cardiomyocyte-specific Sam68 deletion or overexpression and subjected them to angiotensin II infusion or transverse aortic constriction (TAC) surgery to induce pathological cardiac hypertrophy. Mechanistic studies were performed using RNA-seq, metabolomics, and immunoprecipitation analyses.

**Results:** Sam68 expression was significantly elevated in cardiomyocytes of human myopathic and failing hearts. Deletion of Sam68 in adult mouse cardiomyocytes prevented angiotensin II- and TAC-induced cardiac hypertrophy. Conversely, AAV9-mediated overexpression of Sam68 in cardiomyocyte exacerbated cardiac hypertrophy and failure. RNA-seq and metabolomic analyses showed that Sam68 deficiency led to a marked downregulation of pyruvate dehydrogenase kinase 4 (PDK4), which was associated with enhanced cardiac glucose oxidation and oxidative phosphorylation. Mechanistically, Sam68 directly interacts with STAT3, promoting its phosphorylation and nuclear translocation via Src signaling, thereby enhancing PDK4 transcription. Pharmacological inhibition of the Sam68-Src interaction using a specific peptidomimetic molecule mitigated pathological cardiac hypertrophy by attenuating STAT3 phosphorylation and restoring glucose oxidation. Additionally, the Sam68/STAT3/PDK4 signaling axis was significantly unregulated in patients with heart failure.

**Conclusions:** Our findings reveal a novel role of Sam68 in regulating cardiac glucose oxidation, highlighting the potential therapeutic targeting of Sam68 for managing cardiac hypertrophy and heart failure.

**Clinical Perspective What Is New?:**

- Sam68 (Src-associated-in-mitosis-of-68kDa) expression is upregulated in human failing hearts and in mouse models of cardiac hypertrophy and heart failure.
- Deletion of Sam68 in cardiomyocytes mitigates pressure overload-induced cardiac hypertrophy and dysfunction, while overexpression of Sam68 exacerbates these conditions.
- Sam68 exacerbates pressure overload-induced cardiac hypertrophy and dysfunction by binding to STAT3, increasing its phosphorylation and nuclear translocation, which ultimately leads to the upregulation of PDK4 and impairment of glucose oxidation.

**What Are the Clinical Implications?:**

- Upregulation of the Sam68/STAT3/PDK4 signaling axis in cardiomyocytes is associated with the development of cardiac hypertrophy and heart failure.
- Pharmacological inhibition of Sam68 has the potential to improve cardiac energy metabolism, offering new therapeutic options for treating cardiac hypertrophy.
- By modulating the Sam68/STAT3/PDK4 axis, it may be possible to enhance glucose oxidation and mitigate the progression of heart failure.

## Introduction

The heart has a high energy demand, requiring the production of substantial amounts of ATP to sustain its contractile activity^1,2^. The metabolic flexibility of cardiomyocytes allows a healthy heart to adapt its utilization of various metabolic substrates^3^. However, in pathological cardiac hypertrophy, this flexibility is compromised, leading to contractile insufficiency and heart failure (HF)^4^. In healthy individuals, the myocardium derives 60-90% of its ATP from the oxidation of long-chain fatty acids, whereas glucose oxidation contributes to 10-30% of ATP production^2^. During pathological cardiac hypertrophy, there is a metabolic shift from fatty acid oxidation to glycolysis in cardiomyocytes, which helps meet the metabolic demands for hypertrophic growth^5,6^. However, this increased glycolysis is uncoupled from glucose oxidation due to impaired mitochondrial pyruvate uptake, increased pyruvate carboxylation, and reduced activity of pyruvate dehydrogenase (PDH)^7^. Consequently, this uncoupling results in a reduced capacity for ATP generation and an inadequate supply of cardiac energy^8,9^. Moreover, the decoupling of myocardial glycolysis and glucose oxidation leads to the activation of glycolytic bypass pathways, such as the hexosamine biosynthetic pathway (HBP) and the pentose phosphate pathway (PPP), generating metabolites that facilitate cardiac hypertrophy and remodeling^6,10^. Therefore, targeting glucose oxidation may represent a promising strategy to ameliorate cardiac hypertrophy and heart failure.

Pyruvate generated from glycolysis in the heart can either be converted to lactate or transported into mitochondria. Mitochondrial pyruvate is primarily metabolized to acetyl-CoA by PDH, while a smaller fraction is directed towards oxaloacetate^1^. Acetyl-CoA is then catabolized in the TCA cycle, leading to ATP generation. The activity of cardiac PDH is regulated by various isoforms of pyruvate dehydrogenase kinase (PDK1, PDK2 and PDK4) and phosphatases (PDP1 and PDP2), where phosphorylation leads to enzyme inhibition^11^. One of the PDK isoforms, PDK4, is significant upregulated in response to hypertrophic stimuli such as phenylephrine (PE) and angiotensin II (AngII), thereby inhibiting PDH activity in the heart^12^. Previous studies have shown that cardiac-specific PDK4 overexpression impairs metabolic flexibility and exacerbate cardiac hypertrophy under stress stimulation^13^. In contrast, inhibition of PDK4 activity by small molecule compounds, such as dichloroacetate (DCA), can enhance glucose oxidation and improve cardiac hypertrophy and heart function^14–16^. However, the clinical utility of DCA have been limited by its adverse effects on peripheral neuropathy^17^. Therefore, understanding the molecular mechanisms governing PDK4 expression in cardiac hypertrophy is essential for identifying novel pharmacological agents that enhance glucose oxidation and mitochondrial function in HF.

Tyrosine-phosphorylated, Src-associated substrate of 68 kDa protein (Sam68), also known as KH RNA binding domain containing, signal transduction associated 1 (KHDRBS1), is a member of the signal transduction and activation of RNA (STAR) family of proteins. Sam68 plays crucial roles in cell proliferation, differentiation, development, and signal transduction. Additionally, Sam68 is involved in multiple aspects of RNA metabolism, such as transcriptional regulation, alternative splicing modulation, and nuclear localization^18,19^.

Like most RNA-binding proteins in the STAR family, Sam68 contains a STAR domain, a heteronuclear ribonucleoprotein particle K homology (KH) domain, a NK/QUA1 region, and a CK/QUA2 region, which are all essential for its RNA binding activity^19^. In addition to the STAR domain, Sam68 has diverse regions and posttranslational modification sites that contributes to the protein-protein interactions and signal transduction^19^. The proline-rich regions enable Sam68 to bind to the SH3 domains of Src kinases, mediating tyrosine phosphorylation of Sam68 and interaction with SH2 domain-containing proteins^20^. Consequently, Sam68 acts as an adaptor molecule in various signaling pathways^19^. While most studies on Sam68 have focused on its role in RNA binding and cancer^21,22^, recent research has revealed its significant involvement as a metabolic regulator in various pathological processes, including aerobic glycolysis^23^, hepatic gluconeogenesis^24^, adipogenesis^25^, and adipose thermogenesis^26^. However, the role and underlying mechanisms of Sam68 in mitochondrial pyruvate oxidation and cardiac hypertrophy remain poorly understood.

In this study, we provide compelling evidence demonstrating the critical role of Sam68 in impairing myocardial energy metabolism and exacerbating pressure overload-induced cardiac hypertrophy. As an adaptor protein, Sam68 directly binds to STAT3 and facilitates its phosphorylation and nuclear translocation through Src signaling, thereby promoting increased PDK4 expression and reduced pyruvate oxidation, resulting in impaired oxidative metabolism in cardiomyocytes. Genetic or pharmacological inhibition of Sam68 enhances cardiac energy production and mitigates cardiac hypertrophy. Thus, our findings reveal a previously unrecognized role of Sam68 in the regulation of cardiac energy metabolism and highlight the potential therapeutic benefits of inhibiting Sam68 in cardiac hypertrophy and heart failure.

## Methods

The expanded methods are detailed in the Supplementary Materials.

### Human Heart Samples

Left ventricular samples were collected from eleven heart failure patients who underwent heart transplant surgery. Among them, eight were diagnosed with restrictive cardiomyopathy (RCM), and three with hypertrophic cardiomyopathy (HCM). Normal control samples were obtained from healthy donors who died from non-cardiac causes. The study protocol adhered to the Helsinki Declaration and was approved by the Ethics Committee of Union Hospital, Tongji Medical College, Huazhong University of Science and Technology (Approval number: UHCT-IEC-SOP-016-03-01). Written informed consent was obtained from all heart donors and transplant patients.

### Animals

All mice used in this study were subjected to protocols approved by the Institutional Animal Care and Use Committee (IACUC) of Southern University of Science and Technology. Experiments were conducted in 10- to 12-week-old male mice on C57BL/6J background unless otherwise specified. All mice were bred and housed under specific pathogen-free environments at the animal facility of Southern University of Science and Technology, with controlled temperature and humidity and a 12-hour light-dark cycle. Mice were provided with free access to sterilized water and the designated diet. In this study, blinding was implemented to minimize potential biases: the researchers who performed the sample processing and analysis were blinded to the treatment groups.

### Statistical analysis

Data are presented as the mean ± SEM. The Shapiro‒Wilk test was used to assess data normality. The equality of group variances was evaluated using the F test or Brown-Forsythe test. Differences between two groups were assessed using unpaired Student’s *t-*tests when data were normally distributed; otherwise, the nonparametric Mann‒Whitney U test was employed. For comparisons involving more than two groups, one-way ANOVA or Welch’s ANOVA was used, followed by Tukey’s post hoc test (assuming equal variances) or Tamhane’s T2 post hoc test (assuming equal variances). Statistical significance was set at P < 0.05. All statistical analyses were performed using GraphPad Prism 10.2.0 (GraphPad Software, San Diego, CA).

## Results

### Sam68 Is Specifically Upregulated In Cardiomyocytes During Cardiac Hypertrophy and Heart Failure

To investigate the potential association between Sam68 expression and the progression of cardiac hypertrophy and heart failure, we conducted a comprehensive analysis of three bulk RNA-seq datasets (GSE116250, GSE160997 & GSE5406), which included samples from patients with dilated cardiomyopathy (DCM), ischemic cardiomyopathy (ICM), hypertrophic cardiomyopathy (HCM), and normal nonfailing left-ventricular tissues (NF)^27–29^. Our analysis revealed a significant upregulation of KHDRBS1 (Sam68) expression in the myocardium of patients with DCM, ICM, or HCM compared to normal hearts (**Figure 1A-1C**).

**Figure 1.**
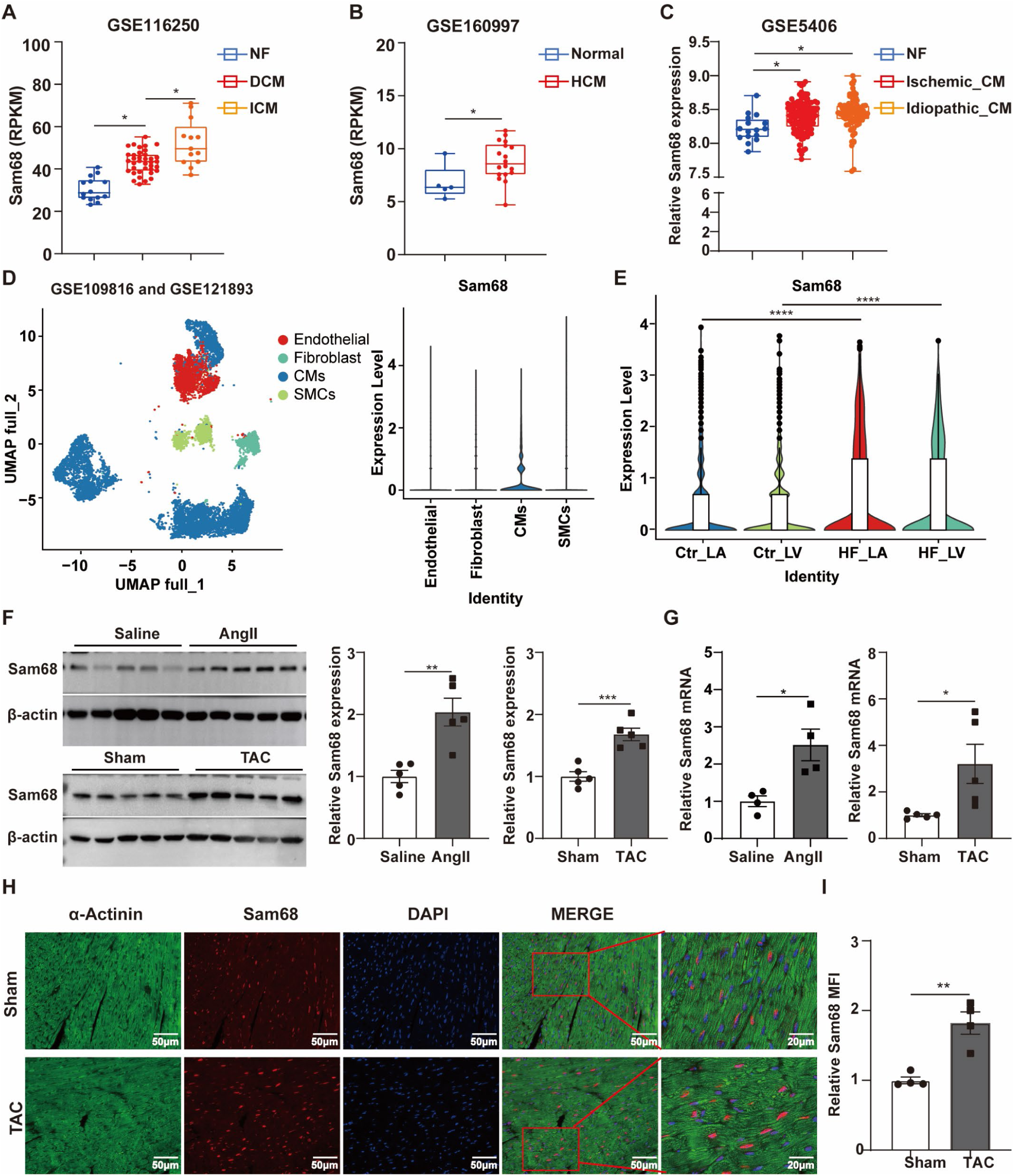
Sam68 expression is elevated in the myopathic and failing hearts. **A-C,** The levels of Sam68 expression in human hearts were quantified from: **A,** RNA-seq data (GSE116250) in patients with non-failing myocardium (NF, n=14), dilated cardiomyopathy (DCM, n=37), or ischemic cardiomyopathy (ICM; n=13); **B,** RNA-seq data (GSE160997) in patients with normal myocardium (Normal; n=5) or hypertrophic cardiomyopathy (HCM; n=18); and **C,** microarray data (GSE5406) in patients with NF (n=16), DCM (n=108), or ICM (n=86). **D-E,** Analyses of single-cell sequencing data (GSE109816 and GSE121893) showing: **D,** major cardiac cell types (*left* panel) and their levels of SAM68 mRNA expression (*right* panel); **E,** Sam68 mRNA levels in cardiomyocytes of left atria and left ventricles from heart failure (HF_LA and HF_LV) and control (Ctr_LA and Ctr_LV) subjects. **F-I,** After receiving continuous Angiotensin II (AngII) infusion for 2 weeks (n=5 for each group) or TAC surgery for 4 weeks (n=5 for each group), mice were analyzed for Sam68 expression in the heart by: **F,** Western blotting (*left* panels, representative blot; *middle* and *right* panels, quantification); **G,** RT-PCR (values were normalized to 18S); **H,** Immunofluorescence co-staining (Sam68 in red, α-Actinin in green, and DAPI in blue); and **I,** Quantification of Sam68 mean fluorescence intensity (MFI) in H (n=4). Data are presented as mean ± SEM, with statistical significance indicated as follows: *p<0.05, **p<0.01, ***p<0.001, ****p<0.0001. Statistical analyses were performed using Student’s t-test (F-H) and one-way ANOVA (A-C and E). RPKM denotes reads per kilobase of transcript per million reads mapped.

To evaluate the expression pattern of Sam68 in cardiac hypertrophy and heart failure at the cellular level, we analyzed single-cell RNA-Seq datasets (GSE109816 & GSE121893) derived from left ventricular (LV) and left atrial (LA) samples obtained from 14 healthy organ donors and 6 heart failure patients. Using molecular profiling, we classified the entire cell population into 4 major types: cardiomyocytes (CMs), endothelial cells (ECs), fibroblasts (FBs), and smooth muscle cells (SMCs) (**Figure S1A and S1B**). We observed that Sam68 expression was markedly enriched in CMs compared to other cell types (**Figure 1D**). Notably, a specific upregulation of Sam68 transcripts was exclusively observed in cardiomyocytes, rather than other cell types, in cardiac tissues obtained from patients with heart failure compared to healthy controls (**Figure 1E and Figure S1C**).

We subsequently assessed the protein and mRNA levels of Sam68 in murine cardiac hypertrophic models induced by continuous AngII infusion and transverse aortic constriction (TAC) surgery, respectively. We demonstrated a significant upregulation of both Sam68 protein and mRNA levels in cardiac tissues following AngII treatment or TAC surgery (**Figure 1F and 1G**). Additionally, an upregulation of Sam68 protein expression was observed in neonatal rat ventricular myocytes (NRVMs) following AngII or phenylephrine (PE) treatment, compared to vehicle treatment (**Figure S1D and S1E**). Moreover, immunofluorescence staining confirmed that Sam68 was predominantly located in the nucleus of cardiomyocytes, and its expression was markedly increased following TAC surgery (**Figure 1H**). Collectively, these findings suggest that Sam68 exhibits specific upregulation within the nuclear compartment of cardiomyocytes under hypertrophic or failing conditions.

### Conditional Cardiomyocyte-Specific Deletion of Sam68 Attenuates AngII-Induced Cardiac Hypertrophy

To investigate the role of Sam68 in cardiomyocytes under both physiological and pathological conditions, we generated a conditional knockout mouse model of Sam68 (Sam68^flox/flox^) by targeting exons 5-8 using our previously established strategies (**Figure S2A**)^24^. We then crossed Sam68^flox/flox^ mice with Myh6^MerCreMer^ mice to generate inducible cardiomyocyte-specific Sam68 conditional knockout mice (genotyped as Sam68^flox/flox^Myh6^MerCreMer^, referred to as Sam68-cKO), whereas Myh6^MerCreMer^ mice were used as littermate controls (referred to as CTR). At 8 weeks of age, both groups were administered tamoxifen intraperitoneally (*i.p.*) at a dose of 20 mg/kg/day for five consecutive days. Two weeks later, we observed a significant reduction in Sam68 protein levels in the hearts of Sam68-cKO mice compared to the littermate controls (CTR) (**Figure S2B**).

To evaluate the impact of cardiomyocyte-specific Sam68 deletion on pressure-overload induced cardiac hypertrophy and heart failure, we administered AngII at a dose of 1.44 mg/kg/day via mini-pump implantation for 14 days. Echocardiography analysis revealed that the dimensions of the interventricular septum (IVS) and left ventricular posterior wall (LVPW) thickness during both diastole and systole were significantly increased in CTR mice after AngII treatment. However, the induction of IVS and LVPW was significantly attenuated in Sam68-cKO mice following AngII treatment (**Figure S3A and Table S1**). Histological analysis showed that Sam68-cKO mice exhibited a decreased cardiac size compared to CTR mice following AngII treatment (**Figure S3B**), along with a reduction in heart weight to body weight (HW/BW) ratio and heart weight to tibia length (HW/TL) ratio (**Figure S3C**). Additionally, Sam68-cKO mice displayed diminished cardiomyocyte size (**Figure S3D**) and improved cardiac fibrosis compared to CTR mice after AngII treatment (**Figure S3E**). Consistent with the improved cardiac performance, both mRNA and protein expression of atrial natriuretic peptide (*Nppa* or ANP) and brain natriuretic peptide (*Nppb* or BNP) were significantly repressed in Sam68-cKO mice compared to CTR mice after AngII treatment (**Figure S3F and S3G**). Collectively, these findings indicate that conditional knockout of Sam68 in adult cardiomyocytes provides effective protection against AngII-induced cardiac hypertrophy.

### Cardiomyocyte-specific Sam68 Deletion in Adult Mice Confers Protection Against Transverse-aortic-constriction (TAC)-Induced Cardiac Dysfunction

Given the improved cardiac performance observed in Sam68-cKO mice following AngII treatment, we next sought to determine the potential protective effects of cardiomyocyte-specific Sam68 deletion against TAC-induced cardiac hypertrophy and dysfunction. Tamoxifen-administered CTR and Sam68-cKO mice were subjected to TAC surgery and monitored for a duration of 4 weeks. Despite a comparable survival rate, we observed a significant reduction in IVS during both diastole and systole in Sam68-cKO mice compared to CTR mice at 2 weeks post-TAC surgery (**Figure S4A, S4B, and Table S2**). However, there were no significant differences in LVPW, ejection fraction (EF), or fraction shortening (FS) in Sam68-cKO mice compared to CTR mice at 2 weeks post-TAC surgery (**Figure S4C, S4D, and Table S2**). Notably, Sam68-cKO mice exhibited restored cardiac systolic function at 4 weeks after surgery, as indicated by decreased LV internal diameter (LVID) along with improved EF and FS compared with CTR mice (**Figure 2A, 2B, and Table S3**). Consistently, histological analysis revealed that Sam68-cKO mice displayed reduced heart size, accompanied by decreased HW/BW and HW/TL ratios compared to CTR mice at 4 weeks post-TAC surgery (**Figure 2C and 2D**). TAC surgery dramatically increased cardiomyocyte size and cardiac fibrosis in CTR mice, whereas these changes were alleviated in hearts of Sam68-cKO mice (**Figure 2E and 2F**). Lastly, we found that the deletion of Sam68 in cardiomyocytes repressed the protein expression of ANP and BNP in heart tissues at 4 weeks post-TAC surgery (**Figure 2G and 2H**). Therefore, these findings suggest that Sam68 knockout in adult cardiomyocytes improves cardiac function by preventing the development of pathological cardiac hypertrophy in response to pressure overload.

**Figure 2.**
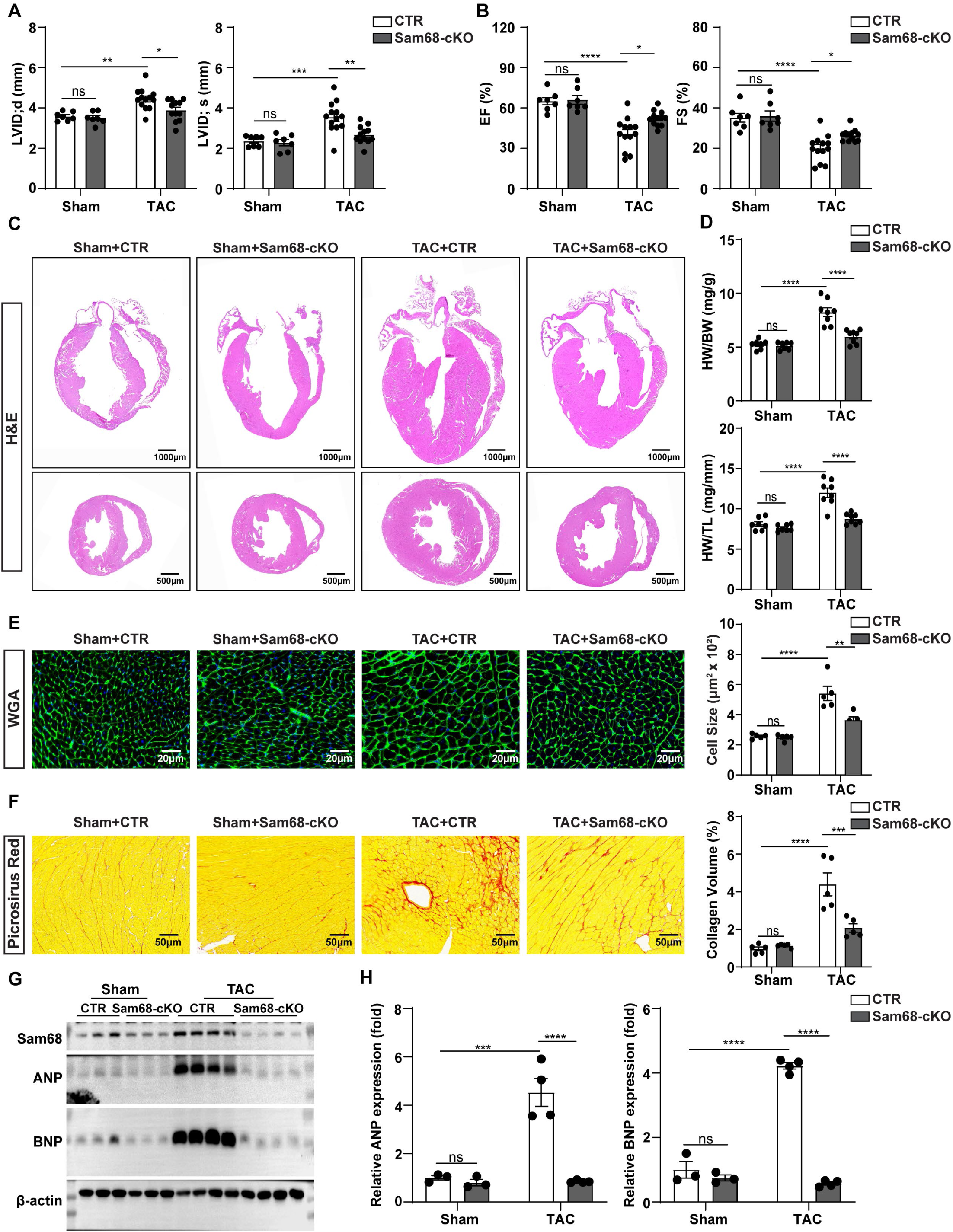
Ablation of Sam68 in adult cardiomyocytes prevents TAC-induced cardiac hypertrophy. Sam68-cKO and control (CTR) mice underwent Sham and TAC surgery and were analyzed 4 weeks later. **A-B,** Echocardiographic analysis of: **A,** Left ventricular (LV) end-diastolic internal dimension (LVID;d; *left* panel) and LV end-systolic internal dimension (LVID;s; *right* panel); and **B,** ejection fraction (EF; *left* panel) and fraction shortening (FS; *right* panel). **C,** Representative hematoxylin-eosin (H&E) staining of longitudinal and cross-sections in the heart. **D,** Quantification of heart weight to body weight (HW/BW) and heart weight to tibia length (HW/TL) ratios. **E,** Representative wheat germ agglutinin (WGA) staining (*left* four panels) and quantification of mean cardiomyocyte cross-sectional area (*right* panel) of ventricular cardiomyocytes. **F,** Representative picrosirius red staining (*left* four panels) and quantification of collagen volume (*right* panel). **G-H,** Western blot analysis (**G**) and quantification of ANP and BNP protein levels (**H**) in the hearts. Data are presented as mean ± SEM, with statistical significance indicated as follows: ns (not significant), *p<0.05, **p<0.01, ***p<0.001, ****p<0.0001. Statistical analyses were performed using two-way ANOVA (A-B, D-F, and H).

### Cardiomyocyte-Specific Sam68 Overexpression Exacerbates TAC-Induced Cardiac Hypertrophy

To investigate the potential effects of Sam68 overexpression on cardiac hypertrophy and heart function, we utilized AAV9 vector with a cardiac troponin T (cTnT) promoter to induce Flag-Sam68 expression in cardiac tissue (referred to as Sam68OE) (**Figure S5A**). Mice receiving AAV9-GFP were used as controls (referred to as GFP). Four weeks after AAV9 injection, successful cardiac-specific overexpression of Sam68 was confirmed by a significant increase in protein levels compared to GFP mice (**Figure S5B**). We then evaluated the impact of cardiac Sam68 overexpression on cardiac hypertrophy and dysfunction following TAC surgery. In contrast to our findings in Sam68-cKO mice, echocardiographic analysis revealed that overexpression of Sam68 in cardiomyocytes exacerbated TAC-induced cardiac hypertrophy at 2 weeks post TAC surgery (**Figure S5C and Table S4**). At 4 weeks after TAC surgery, Sam68 overexpression aggravated cardiac systolic dysfunction and chamber dilation compared to GFP mice (**Figure 3A and Table S5**). Histological analysis showed that Sam68OE mice exhibited increased heart size, HW/BW, and HW/TL ratios compared to GFP mice (**Figure 3B and 3C**). Additionally, Sam68OE mice exacerbated cardiomyocyte enlargement and fibrosis induced by TAC surgery (**Figure 3D and 3E**). Furthermore, both mRNA and protein expression of ANP and BNP were further elevated in the heart of Sam68OE mice compared to GFP mice at 4 weeks post-TAC surgery (**Figure 3F and 3G**). In summary, these data demonstrate that cardiac-specific overexpression of Sam68 exacerbates TAC-induced cardiac hypertrophy and dysfunction.

**Figure 3.**
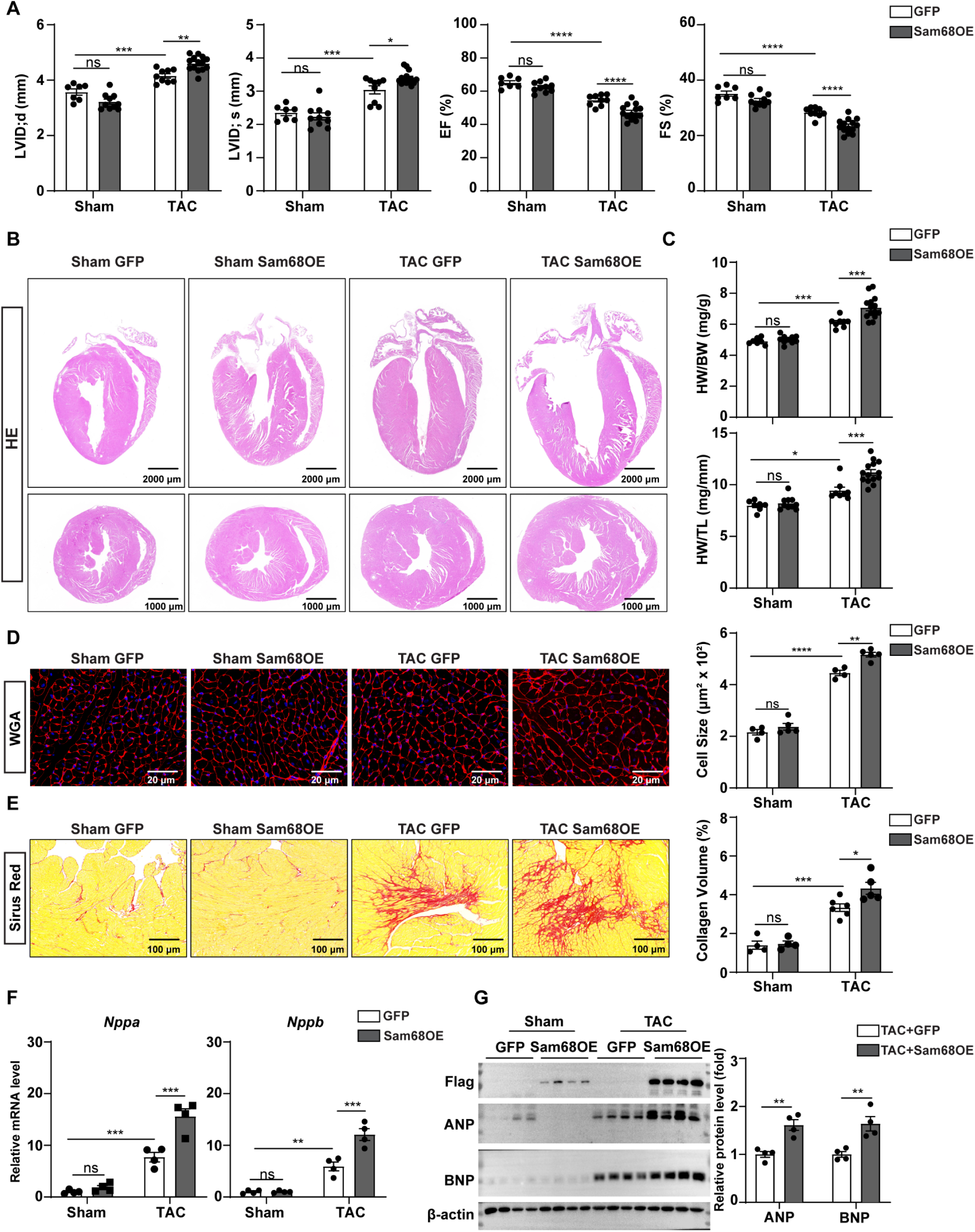
Overexpression of Sam68 in cardiomyocytes exacerbates TAC-induced cardiac hypertrophy. WT mice received *i.p.* injection of AAV9-GFP (GFP) or AAV9-Sam68 (Sam68OE) at a dosage of 5 ×10^11^ genome particles each. The mice then underwent either Sham or TAC surgery, followed by a series of analyses four weeks later. **A,** Echocardiographic analyses of LV end-diastolic internal dimension (LVID;d), end-systolic internal dimension (LVID;s), ejection fraction (EF), and fraction shortening (FS). **B,** Representative images of H&E staining of longitudinal and cross-sections in the hearts. **C,** Quantification of heart weight/body weight (HW/BW) and heart weight/tibia length (HW/TL) ratios. **D,** Representative images of wheat germ agglutinin (WGA) staining (*left* four panels) and quantification of mean cardiomyocyte cross-sectional area (*right* panel) of ventricular cardiomyocytes. **E,** Representative images of picrosirius red staining (*left* four panels) and quantification of collagen volume (*right* panel). **F,** Relative mRNA levels of *Anp* and *Bnp* in the hearts. **G,** Western blot analysis (*left* panel) and quantification of ANP and BNP protein levels (*right* panel) in the hearts. Data are presented as mean ± SEM, with statistical significance indicated as follows: ns (not significant), *p<0.05, **p<0.01, ***p<0.001, ****p<0.0001. Statistical analyses were performed using two-way ANOVA (A, C, and D-F) and Student’s t-test (G).

### Sam68 Promotes Cardiac Hypertrophy by Impairing Oxidative Metabolism

To investigate the mechanisms through which Sam68 promotes cardiac hypertrophy, we performed RNA sequencing on cardiac apical tissues from 1-week AngII-treated Sam68-cKO and CTR mice. We evaluated the impact of Sam68 knockout in cardiomyocytes on the cardiac transcriptome and identified 806 upregulated and 771 downregulated differentially expressed genes (with a P value < 0.05). Subsequently, KEGG pathway analysis revealed that the downregulated genes in Sam68-cKO mice were significantly enriched in pathways associated with cardiac hypertrophy, while the upregulated genes were highly enriched in oxidative phosphorylation (**Figure 4A and 4B**). Consistently, oxidative phosphorylation and mitochondrial metabolic pathways also emerged as the most significantly enriched Gene Ontology (GO) pathways (**Figure 4C**). Furthermore, differential gene analysis revealed a downregulation of genes associated with cardiac hypertrophy in Sam68-cKO hearts (**Figure 4D**), while simultaneously exhibiting a sustained upregulation of genes related to the mitochondrial respiratory electron transport chain (ETC) (**Figure 4E**). To further verify these results, we assessed the protein expression of ETC complex subunits *in vivo*. As expected, the protein expression of ETC complex I (NDUFB8) and complex II (SDHB) were significantly upregulated in the heart of Sam68-cKO mice compared to those of CTR mice after 2 weeks of AngII treatment (**Figure 4F**). Conversely, the expression of ETC complexes (Com I-V) was further repressed in Sam68OE hearts compared with GFP hearts after TAC surgery (**Figure 4G**). Collectively, these findings suggest that Sam68 deletion in cardiomyocytes alleviates cardiac hypertrophy by augmenting mitochondrial oxidative metabolism.

**Figure 4.**
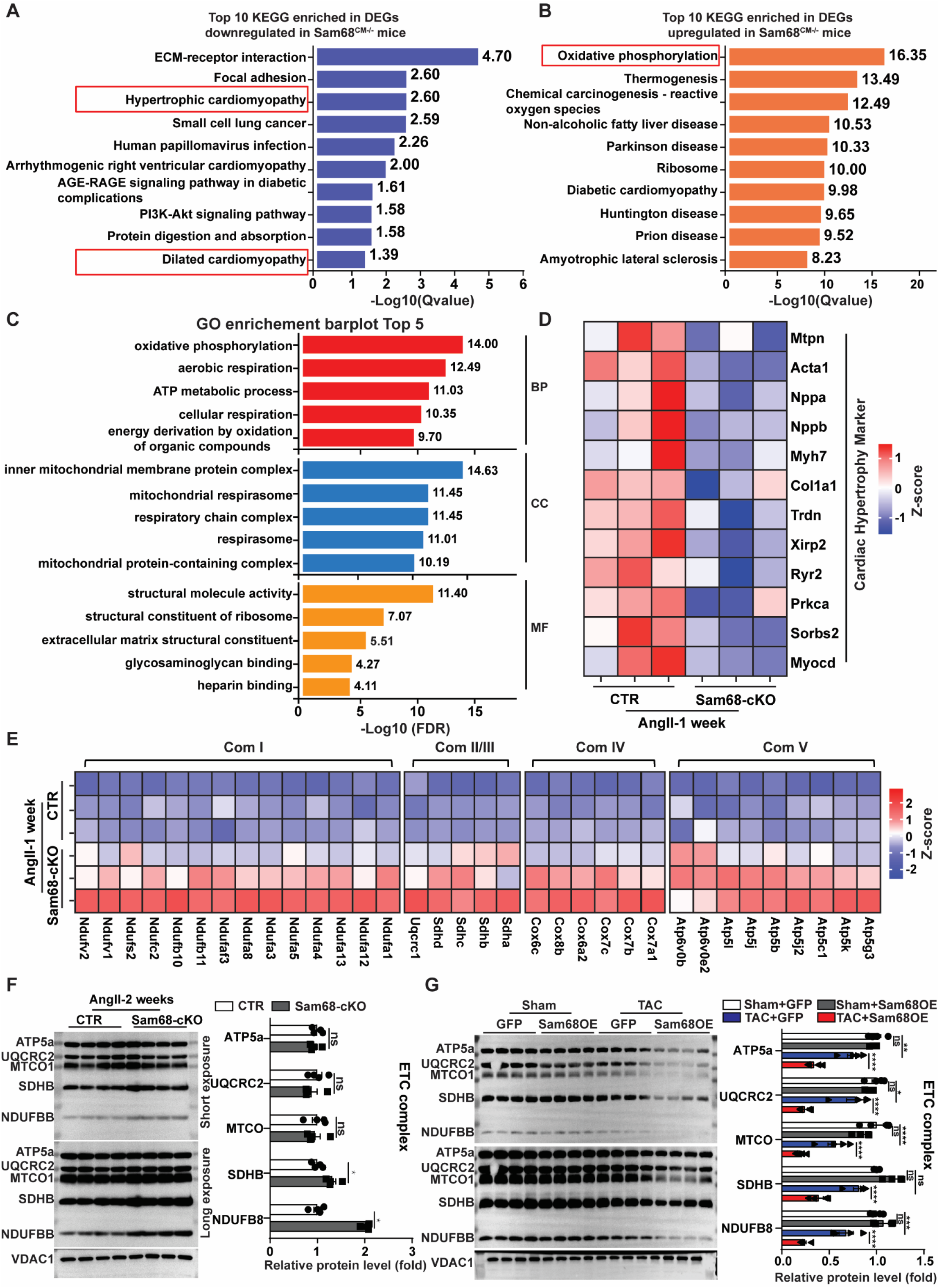
Ablation of Sam68 in cardiomyocytes improves cardiac oxidative metabolism under hypertrophic stimulation. **A-E,** Sam68-cKO and control (CTR) mice were continuously infused with AngII (1.44 mg/kg/day) for 1 week (**A-E**). The transcriptomes in the hearts were analyzed using RNA-seq and compared between the two groups. **A-B,** KEGG enrichment analysis was performed to identify upregulated (**A**) and downregulated (**B**) genes in Sam68-cKO compared to CTR mice. **C,** Go enrichment analysis was conducted to identify differentially expressed genes between the two groups. **D-E,** Heatmaps showing differentially expressed hypertrophy-related genes (**D**) and mitochondrial electron transport chain (ETC) complex I-V genes in Sam68-cKO mice compared to CTR mice. **F,** Western blot analysis (*left* panel) and quantification (*right* panel) of ETC complex subunits were performed in the hearts of Sam68-cKO and CTR mice after 2 weeks of AngII (1.44 mg/kg/day) infusion. **G,** WT mice were injected with AAV9-GFP (GFP) or AAV9-Sam68 (Sam68OE) and subjected to Sham or TAC surgery 3 weeks after injection; the mice were analyzed 4 weeks later by Western blotting (*left* panel) and quantification (*right* panel) of ETC complex proteins. Data are presented as mean ± SEM, with statistical significance indicated as follows: ns (not significant), *p<0.05, **p<0.01,***p<0.001,****p<0.0001. Statistical analyses were performed using Student’s t-test (F) and two-way ANOVA (G).

### Sam68 Deficiency Enhances Glucose Oxidation During Cardiac Hypertrophy

Given the pivotal role of mitochondrial homeostasis in the progression of cardiac hypertrophy, we next investigated the underlying mechanisms through which Sam68 mediates impairment of cardiac oxidative metabolism in response to pressure overload. By conducting a comprehensive analysis of our RNA-Seq data obtained from heart tissues of Sam68-cKO and CTR mice at 1 week post-AngII treatment, we identified 31 upregulated and 64 downregulated differentially expressed genes (with a log2-fold change > 1 and P value < 0.05) (**Figure S6A**). Notably, the expression of *Pdk4*, a gene that encodes a protein regulating pyruvate metabolism and oxidative phosphorylation by directly phosphorylating the E1α subunit of PDH^30^, exhibited prominent downregulation in Sam68-cKO hearts (**Figure S6A**). In the single-cell RNA-seq datasets obtained from heart failure patients, we observed a positive correlation between the expression of PDK4 and Sam68 in cardiomyocytes (**Figure S6B**). Additionally, this positive correlation between *Sam68* and *Pdk4* expression was also verified in the RNA-seq data of cardiomyocytes obtained from mice subjected to TAC surgery (**Figure S6C**). Moreover, a broad upregulation of pyruvate proximal genes was also identified in Sam68-cKO hearts from our RNA-seq data (**Figure S6D**). Consistently, the protein expression of PDK4 and p-PDHα1/PDHα1 ratio were significant downregulated in the hearts of Sam68-cKO mice compared to CTR mice at 2 weeks post-AngII treatment (**Figure 5A**). Conversely, in the hearts of mice subjected to TAC treatment, Sam68OE mice exhibited an elevated protein level of PDK4 and a higher p-PDHα1/PDHα1 ratio compared to GFP mice (**Figure 5B**). These data suggest that Sam68 impairs cardiac oxidative metabolism by upregulating PDK4 expression and inhibition of pyruvate oxidation.

**Figure 5.**
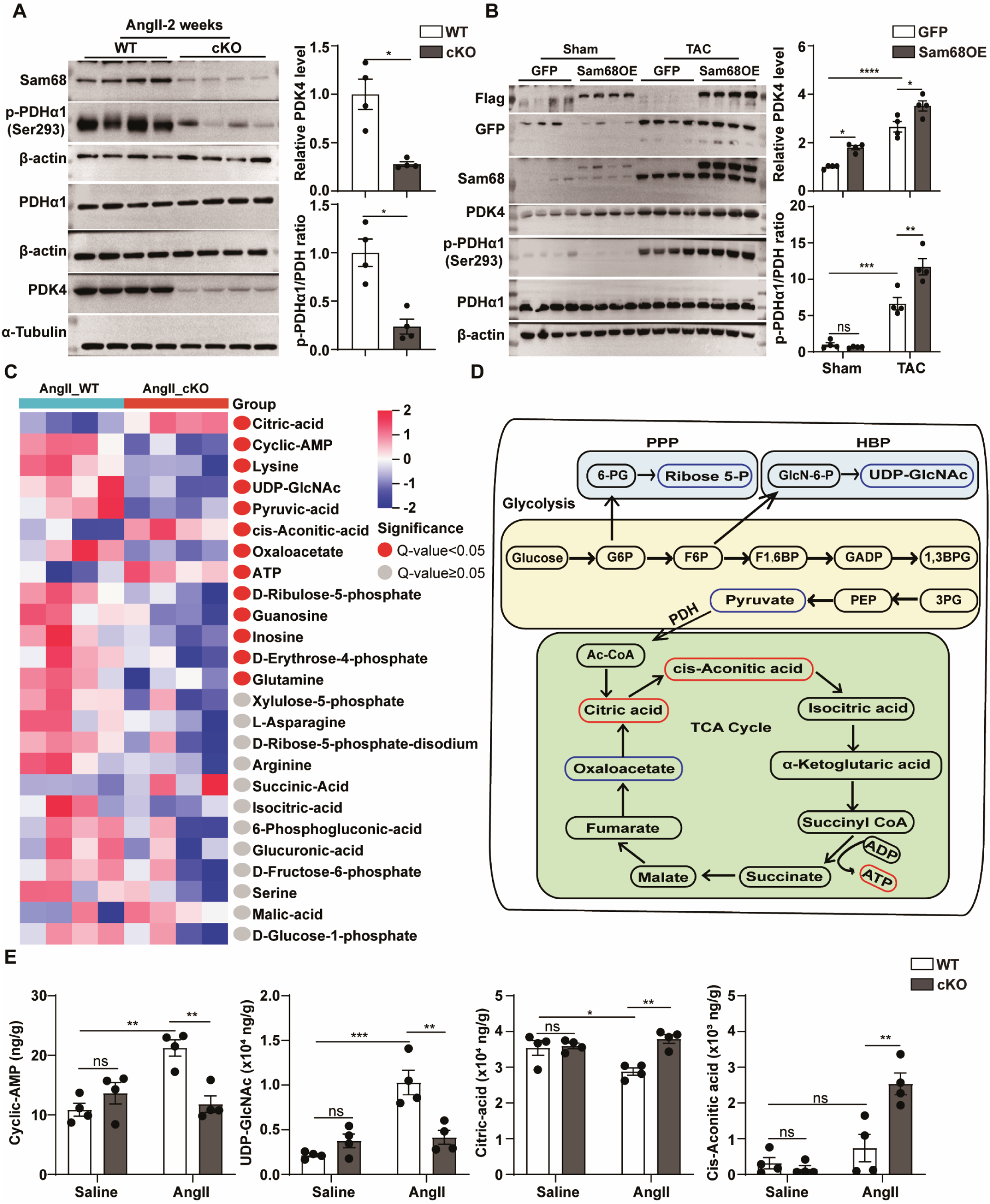
Ablation of Sam68 increases myocardial energy production by promoting pyruvate oxidation. **A-B,** Western blotting analysis and quantification of PDK4, p-PDHα1(Ser293), and PDH expression in the hearts of Sam68-cKO and control (CTR) mice after 2 weeks of continuous AngII infusion for 2 weeks (**A**) and in the hearts of AAV9-GFP (GFP) or AAV9-Sam68 (Sam68OE) transduced mice at 4 weeks after Sham or TAC surgery (**B**). **C-E,** Targeted energy metabolomics analyses were performed in the hearts of Sam68-cKO and CTR mice treated with AngII infusion for 2 weeks. The altered metabolites (Variable importance in projection; VIP>1) in Sam68-cKO mice compared to CTR mice (**C**) are enriched in glycolysis and the TCA cycle (**D;** red boxes, significantly upregulated; blue boxes, significantly downregulated). Key altered metabolites, including Lysine, Cyclic-AMP, UDP-GlcNAc, Ac-CoA, Citric acid, and ATP, were quantified (**E**). Data are presented as mean ± SEM, with statistical significance indicated as follows: ns (not significant), *p<0.05, **p<0.01, ***p<0.001, ****p<0.0001. Statistical analyses were performed using Student’s t-test (A) and two-way ANOVA (B and E).

### Metabolome Analysis Shows That Conditional Knockout of Sam68 in Cardiomyocytes Improves Myocardial Energy Production Under Hypertrophic Stimulation

To further explore the role of Sam68 in the regulation of cardiac energy metabolism under pressure-overload conditions, we conducted targeted metabolomics analyses. A total of 56 metabolites were identified in the hearts of CTR and Sam68-cKO mice at 2 weeks after saline or AngII treatment. These metabolites were further validated using orthogonal partial least squares-discriminant analysis (OPLS-DA), revealing statistically significant differences among the four groups, with samples falling within a 95% confidence interval (Cl) (**Figure S7A**). Differential metabolite analysis and volcano plots, along with a visual heat map depicting all detected metabolites, were generated for the hearts of CTR and Sam68-cKO mice following saline or AngII treatment (**Figure S7B-S7D**). Compared to saline treatment, AngII treatment for 2 weeks significantly upregulated 15 metabolites and downregulated 2 metabolites (VIP>1, P<0.05, and |log2FC|>0.5) in CTR mice (**Figure S7B**). In contrast, after 2 weeks of AngII treatment, Sam68-cKO mice exhibited 2 significantly upregulated and 6 downregulated metabolites in their hearts compared to CTR mice (**Figure S7C**).

Further analysis of the differentially expressed metabolites in the heats from CTR and Sam68-cKO mice following AngII administration revealed that Sam68 deletion in cardiomyocytes resulted in a reduction in pyruvate levels, as well as decreased levels of metabolites associated with the pentose phosphate pathway (ribose-5-P and erythrose-4-P) and hexosamine biosynthesis pathway (UDP-GlcNAc), while concurrently increasing the expression of metabolites related to the tricarboxylic acid cycle (Citric-acid, cis-Aconitic-acid, and ATP) (**Figure 5C**). These findings suggest that deletion of Sam68 enhances the coupling of glycolysis and the TCA cycle, promoting more efficient ATP production and inhibiting glucose metabolism flux into accessory metabolic pathways (**Figure 5D**). Consistently, statistical analysis revealed a significant upregulation of cardiac hypertrophy-related metabolites in the hearts of CTR mice following AngII treatment, including lysine, cyclic-AMP, and UDP-GlcNAc; however, this effect was attenuated in Sam68-cKO mice (**Figure 5E**). Furthermore, cardiomyocyte-specific Sam68 deletion restored TCA cycle activity and cardiac energy production after AngII treatment, resulting in increased levels of citric acid and cis-Aconitic acid compared to CTR mice (**Figure 5E**).

### Sam68 Facilitates STAT3 Phosphorylation and Nuclear Translocation via Src Signaling Transduction

To further elucidate the mechanisms by which Sam68 promotes *pdk4* transcription, we utilized an online tool STRING^31^ to predict interactors associated with Sam68. In addition to previously reported interactors, our analysis revealed a potential interaction between Sam68 and STAT3, a key transcription factor for *Pdk4*^32^ (**Figure S8A**). The binding mode between Sam68 and STAT3 was further determined using ZDOCK 3.0.2 predictions, which showed a direct interaction between the NK domain of Sam68 and the SH2 domain of STAT3 (**Figure 6A**).

**Figure 6.**
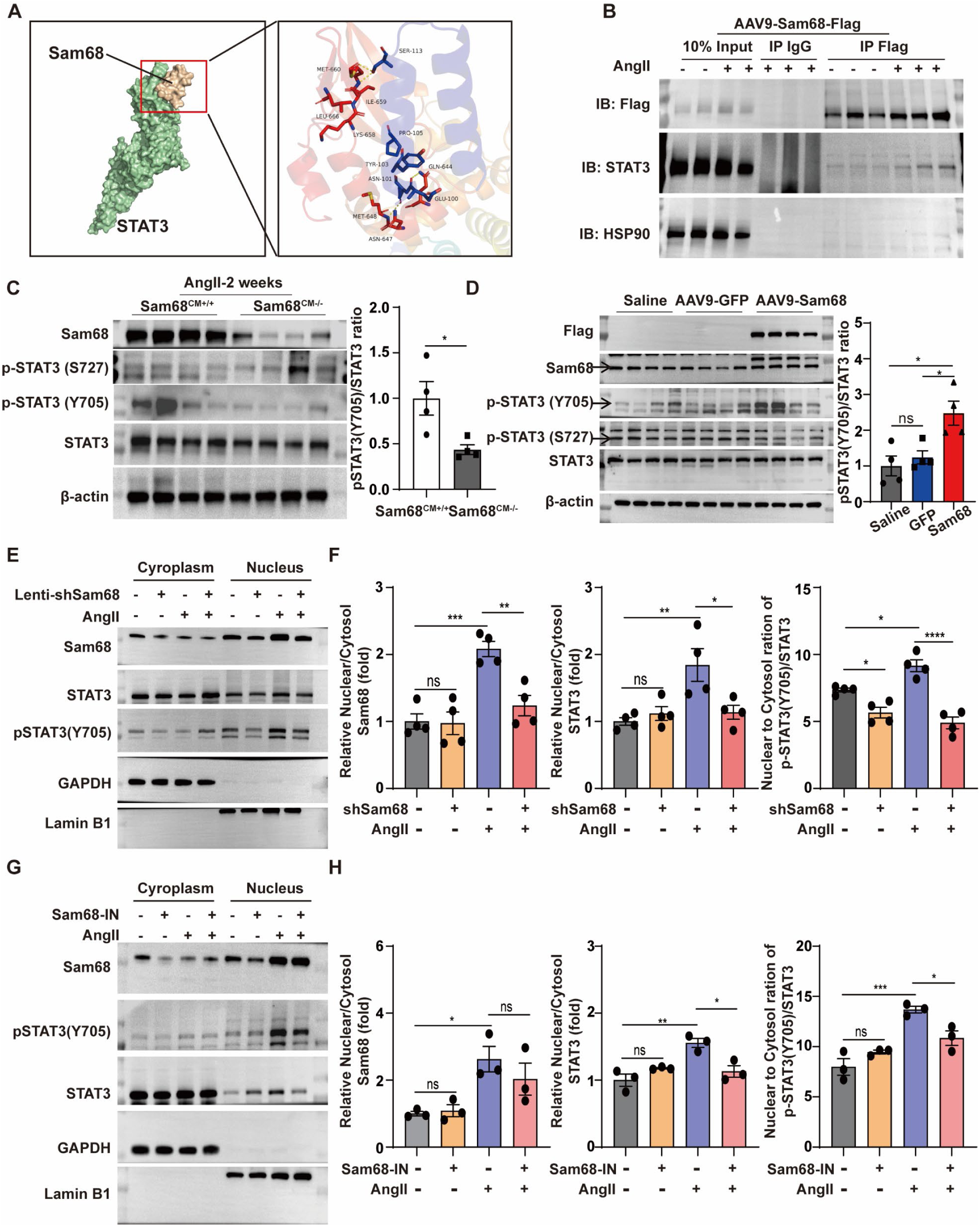
Sam68 interacts with STAT3 and promotes its phosphorylation and nuclear accumulation via Src signaling. **A**, Predicted binding mode of Sam68 and STAT3 based on docking analysis. **B,** Heart lysates from AAV9-Sam68 injected mice, treated with Saline (−) or AngII (+), were subjected to co-IP with anti-FLAG antibodies. The expression of STAT3 in the precipitates was analyzed by immunoblotting. **C,** Western blotting analysis and quantification of p-STAT3 (S727), p-STAT3 (Y705), and STAT3 proteins in the hearts of Sam68-cKO or control (CTR) mice after 2 weeks of AngII infusion. **D,** Western blotting analysis and quantification of p-STAT3 (S727), p-STAT3 (Y705), and STAT3 proteins in the hearts of PBS, AAV9-GFP, or AAV9-Sam68 treated mice. **E-F,** NRVMs were infected with lentiviral Sam68-shRNA vector (+) or none targeting control vector (−) and 24 h later, treated with AngII (1 μM) for 24 h. The protein levels of Sam68, p-STAT3 (Y705), and STAT3 were analyzed in the cytoplasm and nucleus by Western blotting (**E**), and the nuclear-to-cytoplasmic ratios of Sam68 and STAT3 and the p-STAT3-to-STAT3 ratio were quantified (**F**). **G-H,** NRVMs were treated with Vehicle or YB-0158 together with or without AngII (1 μM) for 24 h. The expression of Sam68, p-STAT3 (Y705), and STAT3 were analyzed in the cytoplasm and nucleus by western blotting, and the ratio of nuclear-to-cytoplasmic Sam68 and STAT3 and the p-STAT3-to-STAT3 ratio were quantified. Data are presented as mean ± SEM, with statistical significance indicated as follows: ns (not significant), *p<0.05, **p<0.01, ***p<0.001, ****p<0.0001. Statistical analyses were performed using Student’s t-test (A) and one-way ANOVA (B, F, and H).

We validated the interaction using co-immunoprecipitation assays in the hearts of Sam68OE mice. AngII treatment significantly increased the expression of Flag-Sam68 and enhanced its co-immunoprecipitation with STAT3 (**Figure 6B**). Similarly, we identified the interaction between endogenous Sam68 and STAT3 in NRVMs and demonstrated that AngII treatment increased the co-immunoprecipitation of endogenous Sam68 and STAT3 (**Figure S8B)**. Given the essential role of the SH2 domain in STAT3’s transcriptional activity, we next examined the expression of STAT3-related genes^33^ in our RNA-seq data and found consistent downregulation of these genes in Sam68-cKO mouse hearts compared to CTR mice following 1-week AngII treatment (**Figure S8C**). These data suggest that the interaction between Sam68 and STAT3 contributes to STAT3’s transactivity.

Canonical phosphorylation of STAT3 at Tyr705 results in its dimerization, nuclear translocation, and transactivation^34^. We found that p-STAT3 (Y705) levels were decreased in the hearts of Sam68-cKO mice compared to CTR mice after 2 weeks of AngII treatment (**Figure 6C)**. Conversely, overexpression of Sam68 significantly increased p-STAT3(Y705) levels under basal conditions (**Figure 6D**). Considering the crucial role of STAT3 phosphorylation in facilitating its nuclear translocation, we investigated the subcellular localization of p-STAT3(Y705) and STAT3 in NRVMs following AngII treatment, both with and without Sam68 knockdown. Importantly, AngII treatment enhanced the nuclear accumulation of Sam68, STAT3, and p-STAT3 (Y705), effects that were attenuated upon Sam68 knockdown (**Figure 6E and 6F**).

Sam68 is widely recognized as a direct substrate of Src protein and function as a docking protein for signal transduction^35^. YB-0158 is a reverse-turn peptidomimetic small molecule that binds to Sam68 with high affinity and directly disrupts its interaction with SH3 domain-containing proteins, such as Src^36,37^. To investigate whether Sam68 promotes STAT3 transcriptional activation via Src signaling, we co-treated NRVMs with AngII and YB-0158. Remarkably, inhibition of the Src-Sam68 interaction by YB-0158 effectively prevented AngII-induced nuclear accumulation of STAT3 and p-STAT3(Y705) (**Figure 6G and 6H**). Collectively, these findings suggest that hypertrophic stimuli enhance the interaction between Sam68 and STAT3, facilitating STAT3 phosphorylation and subsequent nuclear accumulation. Furthermore, this mechanism governs STAT3 transcriptional activation via Src signaling transduction.

### Therapeutic Implications of Sam68 Inhibition in Cardiac Hypertrophy

Given the essential role of the Sam68/STAT3/PDK4 axis in regulating mitochondrial glucose oxidation and cardiac hypertrophy, we investigated whether inhibiting Sam68 function could exert protective effects against cardiac hypertrophy and provide potential clinical benefits. We first assessed the effects of YB-0158 on AngII-induced cardiac hypertrophy *in vivo* (**Figure S9A**). YB-0158 treatment significantly attenuated AngII-induced cardiac hypertrophy, as evidenced by reduced IVS dimensions (**Figure S9B and Table S6**), decreased HW/BW and HW/TL ratios (**Figure S9C**), and reduced cardiac size (**Figure S9D**). Additionally, YB-0158-treated mice exhibited smaller cardiomyocytes and less cardiac fibrosis following AngII treatment (**Figure S9E-S9G**). Moreover, protein levels of p-STAT3(Y705), p-PDHα1(S293), and ANP were decreased in the hearts of YB-0158-treated mice compared to vehicle-treated mice following AngII administration (**Figure S9H and S9I**). These findings suggest that inhibiting Sam68 ameliorates AngII-induced cardiac hypertrophy.

To further explore the therapeutic effects of YB-0158 on TAC-induced cardiac hypertrophy and dysfunction, C57BL/6J mice underwent Sham or TAC surgery and were pretreated with YB-0158 (*i.p.* 5 mg/kg/day) or vehicle control for three days before surgery and seven days following surgery (**Figure 7A**). Four weeks post-surgery, echocardiographic analysis revealed that YB-0158 treatment mildly affected ventricular wall thickness, but significantly reduced LV end-systolic diameter (LVID,s) compared to vehicle-treated mice (**Figure S10A, S10B, and Table S7**). Cardiac systolic function was also improved in YB-0158-treated mice, as evidenced by increased ejection fraction (EF) and fractional shortening (FS) (**Figure 7B**). Histological analysis showed that YB-0158 significantly mitigated TAC-induced cardiac hypertrophy, reducing HW/BW (**Figure S10C and S10D**) and HW/TL (**Figure 7C and 7D**) ratios, as well as attenuating cardiomyocyte size and fibrosis (**Figure 7E and 7F**). Furthermore, improved cardiac performance in YB-0158-treated mice after TAC surgery was consistent with decreased ANP and BNP protein levels compared to the vehicle-treated group (**Figure 7G**). Importantly, YB-0158 tended to restore impaired glucose oxidation following TAC surgery, as evidenced by reduced levels of PDK4, p-PDHα1(S293), and p-STAT3(Y705) (**Figure 7H**). Collectively, these findings suggest that pharmacological inhibition of Sam68 with YB-0158 demonstrates functional and structural benefits in mitigating cardiac hypertrophy and dysfunction.

**Figure 7.**
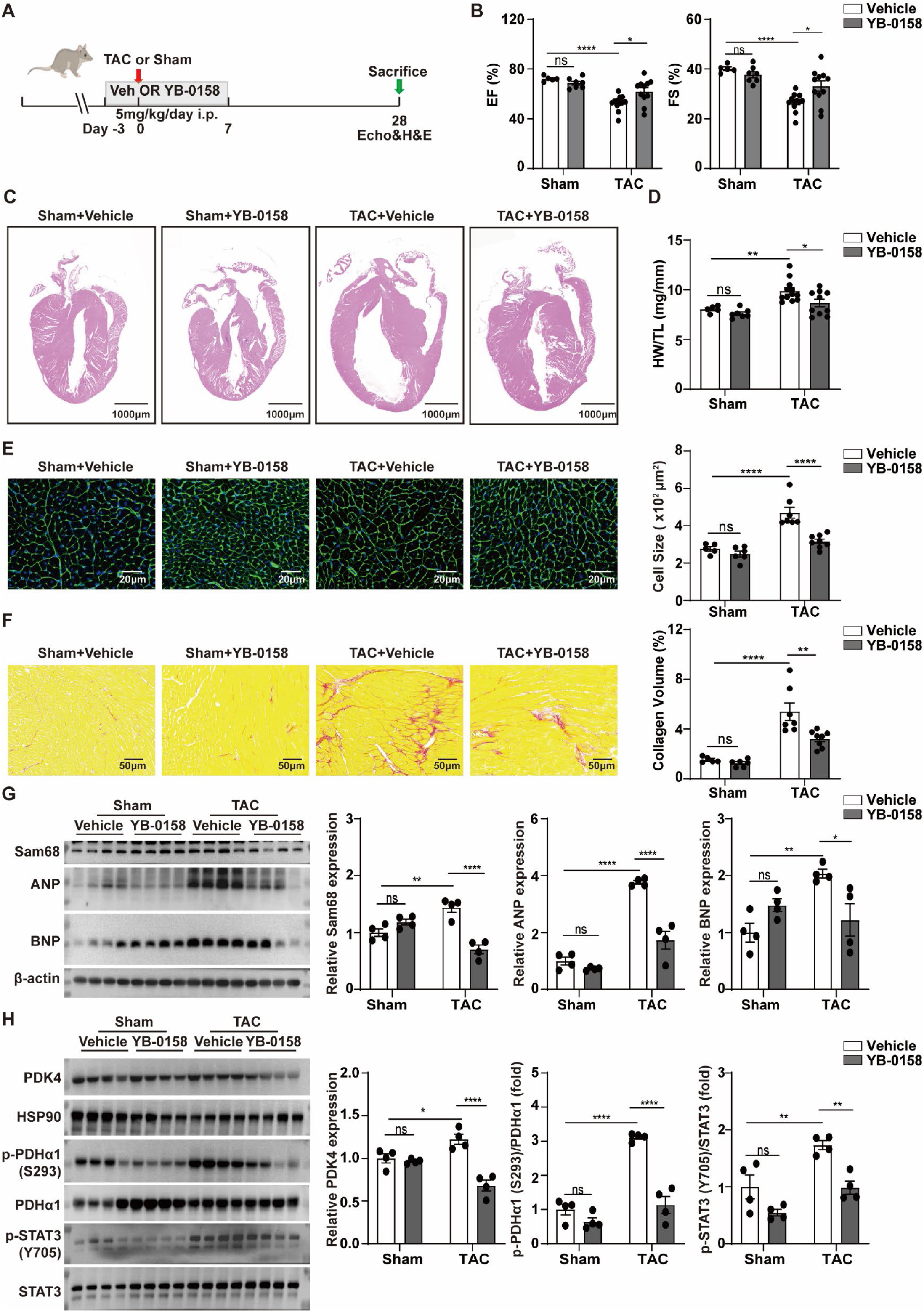
Pharmacological inhibition of Sam68 prevents TAC-induced cardiac hypertrophy. **A,** Schematic diagram of the experimental design for Sam68 inhibition. C57BL/6J mice were *i.p.* injected daily with YB-0158 or vehicle for 3 d, then subjected to TAC or Sham surgery. YB-0158 or vehicle injections continued for another week, and mice were observed for 4 weeks before assessments. **B,** Echocardiographic analysis of ejection fraction (EF) and fraction shortening (FS). **C,** Representative H&E staining of heart longitudinal sections. **D,** Quantification of heart weight/tibia length (HW/TL) ratios. **E,** Representative images of wheat germ agglutinin (WGA) staining (*left* four panels) of the hearts and quantification of ventricular cardiomyocyte cross-sectional area (*right* panel). **F,** Representative images of picrosirius red staining (*left* panel) of the hearts and quantification of collagen volume (*right* three panels). **G,** Western blotting analysis and quantification of Sam68, ANP, and BNP protein levels in the hearts. **H,** Western blotting analysis and quantification of the PDK4 protein level and the ratios of p-PDHα1(S293)/PDHα1 and p-STAT3(Y705)/STAT3 in the hearts. Data are presented as mean ± SEM, with statistical significance indicated as follows: ns (not significant), *p<0.05, **p<0.01, ****p<0.0001. Statistical analyses were performed using two-way ANOVA (B, D-H).

### Upregulation of the Sam68/STAT3/PDK4 Signaling in Heart Failure Patients

To further establish the clinical relevance of the Sam68/STAT3/PDK4 axis in heart failure, we examined ventricular heart specimens from heart failure patients and healthy donors (**Table S8**). As shown in **Figure 8A** and **8B**, the levels of Sam68, p-STAT3 (Y705)/STAT3 ratio, PDK4, and p-PDHα1(S293)/PDHα were significantly upregulated in patients with heart failure compared to control donors, concurrent with increased ANP protein expression. These results suggest that the Sam68/STAT3/PDK4 axis is aberrantly upregulated in heart failure patients, impairing cardiac glucose oxidation in the clinical setting.

**Figure 8.**
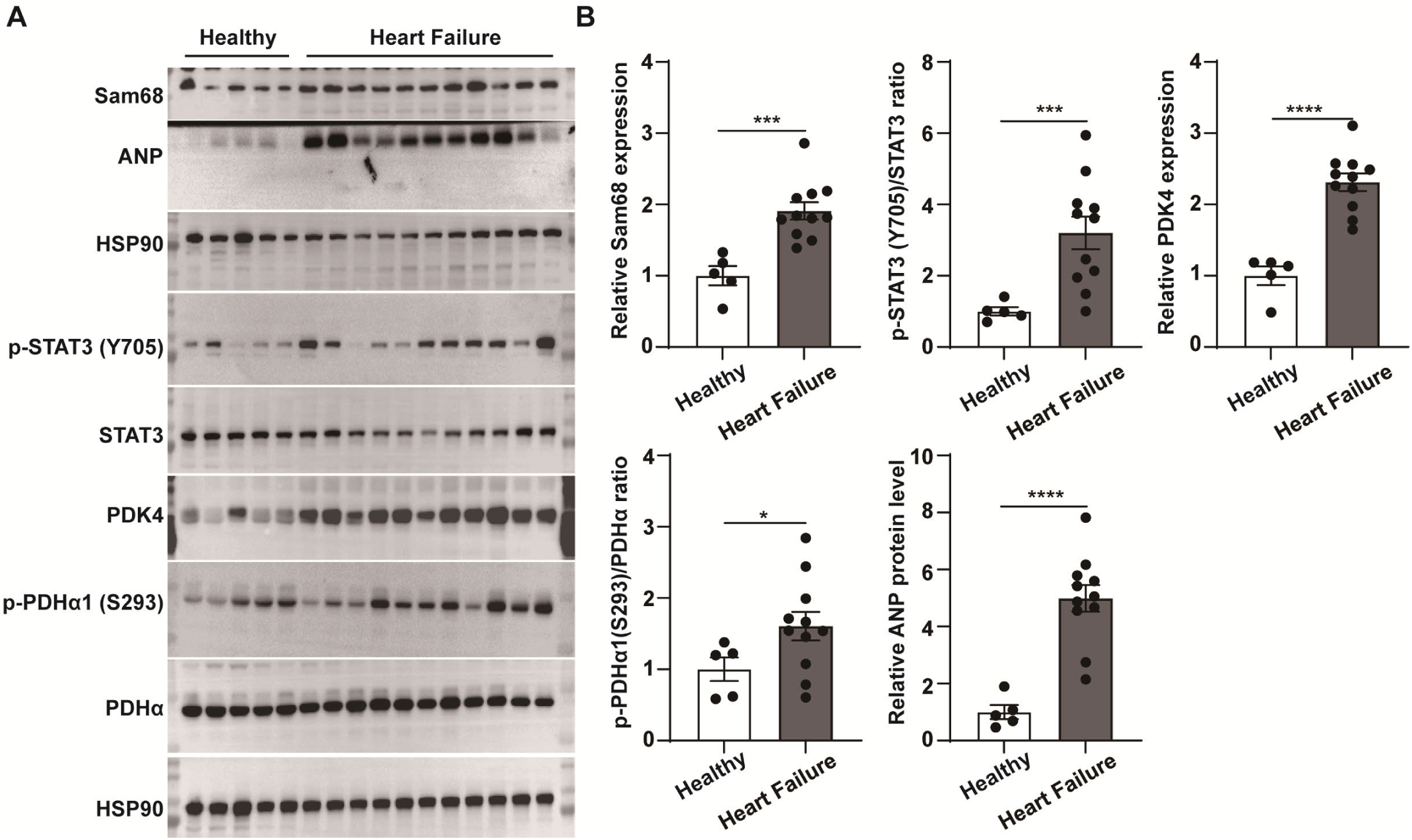
Sam68/STAT3/PDK4 signaling is upregulated in failing human hearts. **A,** Western blotting analysis of Sam68, p-STAT3 (Y705), STAT3, p-PDHα1(S293), PDHα, PDK4, and ANP proteins in human left ventricular specimens obtained from healthy donors (NF) or patients with heart failure (HF). **B,** Quantification of the Sam68 and PDK4 protein levels, as well as the p-STAT3 (Y705)/STAT3 and p-PDHα1(S293)/PDHα ratios. Data are presented as mean ± SEM, with statistical significance indicated as follows: *p<0.05, ***p<0.001, ****p<0.0001. Statistical analyses were performed using Student’s t-test.

## Discussion

Alterations in cardiac energy metabolism during pathological cardiac hypertrophy exacerbates contractile dysfunction and heart failure^38^. Despite numerous studies demonstrating the impairment of cardiac glucose oxidation in response to hypertrophic stimuli, contributing to myocardial energy deficit and heart failure, the underlying mechanism remains elusive^1,3^. In this study, we provide compelling evidence that Sam68 is markedly upregulated in failing human myocardium and hypertrophic murine hearts, exerting detrimental effects on cardiac glucose oxidation and function. Deletion of Sam68 in cardiomyocytes effectively improves glucose oxidation and cardiac oxidative phosphorylation, alleviating cardiac hypertrophy and dysfunction. Conversely, augmenting Sam68 abundance impairs glucose oxidation and exacerbates TAC-induced cardiac hypertrophy. Mechanistically, upon exposure to hypertrophic stimuli, Sam68 promotes the upregulation of PDK4 expression by directly interacting with STAT3, a transcription factor governing PDK4. The interaction between Sam68 and STAT3 induces phosphorylation of STAT3 and its subsequent nuclear accumulation, thereby enhancing the transcriptional activity of STAT3. Furthermore, we demonstrate that inhibition of Sam68 effectively attenuates the development of pathological cardiac hypertrophy by suppressing STAT3 phosphorylation and reducing PDK4 expression. In clinical setting, the Sam68/STAT3/PDK4 signaling pathway exhibited aberrant upregulation in heart failure patients compared to control donors. Our findings expand the understanding of altered energy metabolism in cardiac hypertrophy and provide a promising therapeutic target for reversing impaired cardiac glucose oxidation in the treatment of cardiac hypertrophy and heart failure (**Figure S11**).

During pathological cardiac hypertrophy, cardiac metabolic reprogramming is characterized by a shift of myocardial energy substrates from fatty acid oxidation (FAO) to glycolysis^1,2^. Although initially considered a compensatory mechanism for a higher rate of energy supply, the increased glycolysis is not accompanied by pyruvate oxidation, resulting in reduced substrate influx into the TCA cycle and a smaller capacity of ATP synthesis^39^. Certain compounds, such as DCA and perhexiline, have been developed to enhance the glucose oxidation and improve cardiac performance, but their clinical application remains limited by unfavorable toxicological profiles and insufficient efficacy in patients^7^. Recently, clinical studies have demonstrated the efficacy and safety of Ninerafaxstat in treating hypertrophic cardiomyopathy patients by switching cardiac metabolism from FAO to glucose oxidation, thereby highlighting the therapeutic potential of cardiac energy regulation^40^. In this study, we have identified Sam68 as a novel therapeutic target by directly inhibiting cardiac pyruvate oxidation in the context of pathological cardiac hypertrophy. A recent study has indicated that Sam68 may be involved in lncRNA Gm15834-induced cardiac hypertrophy, as evidence by *in vitro* overexpression of full-length of Sam68 in HL-1 cardiomyocytes^41^. However, the *in vivo* function and mechanism of Sam68 in cardiac hypertrophy remain largely unknown. After generating cardiac-specific Sam68 knockout and overexpression mice, we have demonstrated that Sam68 suppresses pyruvate oxidation and exacerbates cardiac dysfunction under pressure overload. To the best of our knowledge, this is the first study to report Sam68 as a metabolic regulator of cardiac hypertrophy progression.

PDH E1α is a subunit of the pyruvate dehydrogenase (PDH) complex, which catalyzes the conversion of pyruvate to acetyl-CoA, providing the primary link between glycolysis and the TCA cycle. The main regulators responsible for phosphorylating and inactivating PDH E1α are the serine-specific kinases PDK 1-4, with PDK2 exhibiting the highest phosphorylation activity on Ser293 of PDH E1α, followed by PDK4, PDK1, and then PDK3^42^. In our study, we observed a specific downregulation of the *pdk4* transcript in the hearts of Sam68-cKO mice under AngII treatment, while no significant changes were detected for other PDKs compared to control mice. Furthermore, we discovered a positive correlation between *pdk4* levels and *Sam68* expression after comprehensive analysis of public RNA-seq datasets from heart failure patients and primary cardiomyocytes of TAC-treated mice. This finding is consistent with previous observations that PDK4 is upregulated in hypertrophic mouse hearts^43^. The phenotype observed in Sam68-cKO mice, characterized by reduced cardiomyocyte size and enhanced systolic function under stress stimulation, aligns with findings in PDK4-deficient mice^44^. This suggest that Sam68 exacerbates pathological cardiac hypertrophy following pressure overload by upregulation of PDK4 expression in cardiomyocytes. Beyond its interaction with the PDH complex, PDK4 may also exert effects through other mechanisms. For instance, PDK4 has been shown to bind to and stabilize the cAMP-response element-binding (CREB) protein, leading to the activation of mTORC1^45^. Given the pivotal role of mTORC1 activation in promoting cardiac hypertrophy^46^, future studies may explore the non-PDH-dependent mechanisms of the Sam68-PDK4 axis, particularly its interaction with mTORC1, to fully understand its role in cardiac hypertrophy.

The expression of PDK4 has been shown to be controlled by transcriptional factors FoxO1 and PPARα in cardiomyocytes^47,48^. However, in our study, FoxO1 and PPARα expression showed little difference in the hearts from Sam68-cKO and control mice under pressure overload (data not shown). STAT3 serves as a signaling transducer that mediates the transmission of cytosolic stimuli to the nucleus in response to various pathological stimulation^49^. Activation of STAT3 has been associated with adverse outcomes in cardiac hypertrophy and heart failure^33^. Previous ChIP-seq data and ChIP assays have demonstrated the binding of STAT3 at the promoter region of PDK4, augmenting the transcription of PDK4 and exhibiting an inverse correlation with oxidative phosphorylation^32^. By conducting comprehensive bioinformatics analysis and serial *in vitro* and *in vivo* experiments, we have discovered a direct interaction between the NK domain of Sam68 and the SH2 domain of STAT3. Furthermore, we also present persuasive evidence that Sam68 is indispensable for the phosphorylation and nuclear translocation of STAT3 in response to hypertrophic stimuli. Collectively, these studies substantiate the role of Sam68 in promoting PDK4 transcription through activation of STAT3. The involvement of Sam68 in STAT3 phosphorylation has been previously reported in human peripheral mononuclear cells under leptin stimulation and in TNF-α-induced chondrocytes^50,51^. However, the mechanism is still not clear. By employing small molecules that disrupt the interaction between Sam68 and Src, we demonstrate that Sam68 modulates STAT3 phosphorylation and nuclear translocation via the transduction of Src signaling.

Recent investigations have identified Sam68 as a prominent target for a class of reverse-turn peptidomimetic drugs, which effectively inhibit Wnt/β-catenin mediated transcription^52^. Among these peptidomimetic small molecules, YB-0158 exhibits superior affinity towards Sam68 compared to others and effectively disrupts the Sam68-Src interaction while blocking cancer stem cell activity^36^. In this study, we demonstrate that treatment with YB-0158 significantly improved cardiac hypertrophy under both AngII and TAC treatment, highlighting its therapeutic potential for managing pathological cardiac hypertrophy. Importantly, YB-0158 exhibited no adverse effects on healthy tissues and did not induce any alterations in animal behavioral or clinical parameters, further supporting the safety profile of Sam68 modulation by reverse-turn peptidomimetics^52^. Interestingly, the compound CWP-291 was previously reported as a Sam68-targeting peptidomimetic that has reached phase-1 and 2 clinical trials for multiple myeloma (NTC02426723) and AML (NCT03055286), but its structure is yet undisclosed^53^. Therefore, it would be interesting to investigate whether CWP-291 can prevent cardiac hypertrophy in human patients and how much structural similarity it shares with YB-0158 in further studies.

In conclusion, our study reveals that cardiac Sam68 plays a crucial role in regulating cardiac hypertrophy by activating the STAT3-PDK4 axis. This activation leads to the suppression of glucose oxidation and the impairment of cardiac energy production. These findings not only offer new insights into the regulation of cardiac energy metabolism but also highlight a potential therapeutic strategy for managing cardiac hypertrophy.

## Data Availability

The majority of the data supporting the findings are included in this report. Additional data can be obtained from the corresponding author upon reasonable request.

## Author Contributions

J.A. designed and conducted the experiments, analyzed the data, and wrote the manuscript. C.H. conducted the experiments and revised the manuscript. Y.J., J.S., J.H., and S.X. conducted the experiments and analyzed the data. C.W. performed the bioinformatics analyses. H.L. and N.D. provided key research materials. G.Q. conceived and designed the experiments, analyzed the data, and revised the manuscript. All authors approved the final version of the manuscript.

## Acknowledgements

We would like to express our gratitude to the Gene Denovo Corporation for their support in bioinformatics analysis through their Omicshare Cloud Analysis Platform.

